# Interactive Learning with ChatGPT: Hands-On Practice and Real-Time Feedback in Health Sciences Education for SMART Goal Writing

**DOI:** 10.1101/2024.06.11.24308786

**Authors:** Julie E. Speer, Sara M. Parker, Brittany L. Williams

## Abstract

Physical therapy (PT) students often fail to master documentation skills, such as goal writing, because they struggle to engage in the material early in the curriculum. Therefore, we sought to leverage ChatGPT to create an active learning experience with personalized feedback and hands-on practice. During the activity, students (n=48) learned to use ChatGPT and employed these techniques to learn about goal writing in PT and the SMART (Specific, Measurable, Attainable, Relevant, Time-bound) framework. Next, students engaged in a clarifying lecture before using ChatGPT to generate scenarios for which they drafted or edited SMART goals and asked ChatGPT for feedback. Pre-tests and post-tests were used to measure the impacts of the experience and to capture student perspectives. As a function of the activity, students were better able to recognize the purposes of SMART goals (p<0.0001) and write better goals (p<0.0001). They also showed increased confidence in their abilities (p<0.0001). Furthermore, student responses suggested that they enjoyed the activity (m=3.5/4) and found it helpful (m=3.7/4). Following the activity, 27 students continued to use ChatGPT to practice or study. This activity represents a novel approach for using generative artificial intelligence (AI) in the classroom to help students actively explore the topic of goal writing. Additionally, this modeled responsible use of AI for health care applications. This well-received activity can be easily scaled to include more complex tasks or group discussions, or adapted for use as an asynchronous assignment related to documentation or topics across health science education.

## Background

Clinical documentation is a critical aspect of patient care across health science disciplines^1–4^. It serves as a means of communication and record keeping and because these notes are shared amongst interested parties (e.g., billing specialists, legal representatives, clinicians, researchers, patients), the content must be comprehensive and fulfill multiple requirements^4^. Documentation is an essential component of graduate physical therapy (PT) education, and it is often one of the first classes that students take in in their coursework^5^. Learning clinical documentation supports students in developing critical skills for medical record keeping including those related to patient care delivery, management, and support. In addition, this information is pertinent for billing and reimbursement^3^ and sets a foundation on which students will build throughout their clinical education and training^1,2^. However, students often struggle to learn these skills early in the curriculum, in part, because the content is taught before clinical reasoning has been developed, causing decreased engagement and motivation^6,7^. Additionally, studies have shown that this skill gap can persist into practice, resulting in inaccurate documentation that can have serious ramifications for individual providers and health care systems^8,9^.

Given the importance of these skills and the challenges in teaching them in a way that promotes learner engagement, we sought to leverage emerging educational technologies and contemporary pedagogical approaches. Recent scholarship has identified a role for generative artificial intelligence (AI) and large language model (LLM) tools like ChatGPT (Chat Generative Pre-trained Transformer) both in health care delivery and in education across disciplines^10–19^. These tools have not been publicly available long enough for extensive examinations of the long-term impacts of using ChatGPT or other forms of AI in education.

However, the literature has more broadly begun to illustrate that when aligned intentionally with the learning goals and course context, chatbots like ChatGPT have the potential to improve engagement, increase skill transfer, and promote learning outcomes^14,20,21^. These effects largely derive from the ability of the tool to summarize complex information, generate scenarios, provide feedback, and serve as a virtual tutor. As these strengths of ChatGPT align well with the learning problem previously described, we created and implemented a novel classroom activity that could be completed in under two hours. The design of the activity leveraged AI as an educational tool along with evidence-based active learning techniques and contemporary theories including constructivism^22^ and the ARCS model^23^. Taking these approaches together, we scaffolded the experience in a way that allowed students to independently explore the topic of SMART goals by posing questions to the AI, review the material as a group to check for misunderstandings, and finally to experiment with applying their knowledge to scenarios while getting real-time personalized feedback from the AI and critically evaluating goals written by ChatGPT.

## Methods

### Overview

We developed this learning activity for implementation in week six of a 10-week, in-person, documentation course for first year physical therapy students. Each class session in this course was 90-minutes and the majority of the activity was completed within one session as outlined below. In order to measure the efficacy and impacts of this activity, we utilized a pre/post-test model (approved by ATSU Arizona IRB, #2023-142, exempt protocol). At the start of class (on 8/22/23), we verbally communicated instructions and expectations regarding this activity and the educational research to the students. While all students participated in the activity, students actively chose whether to opt-in (n=48) or opt-out (n=6) of having their responses analyzed towards the research efforts.

### Learning Activity

The activity itself was divided into segments to scaffold the learning (Figure 1), with the majority of components disseminated using the survey platform Qualtrics. Before introducing SMART-goal related content, students first created OpenAI accounts (if they didn’t have one already) and practiced using ChatGPT (version 3.5; August 2023) through what we called the “ChatGPT Playground” (Appendix 1). This was designed as an introduction to the tool and prepared students with the necessary knowledge and skills to use ChatGPT as a virtual tutor in the subsequent segments. Next, the students engaged with instructor-provided prompts to acquire knowledge about the SMART goal framework from ChatGPT (Learning Activity 1; Appendix 1), with ongoing support and troubleshooting assistance readily accessible from the instructor and researchers. During this part of the activity, students asked ChatGPT to tell them about SMART goals and to explain why they are commonly used in healthcare. Additionally, through this dialogue with ChatGPT, students asked for further clarification and examples. Once students had completed this stage, the instructor provided a brief clarifying lecture (Appendix 2) and checked for any misconceptions. In the final segment, students interacted with ChatGPT as a tool for feedback as they wrote, revised, and appraised SMART-goals based on clinical scenarios (Learning Activity 2; Appendix 1). For example, students asked ChatGPT to draft a scenario for them and based on the AI-generated response, they wrote their own SMART-goal. Then they were instructed to ask ChatGPT for feedback on their goal and determine a final revised version. Students also worked through examples where ChatGPT generated the SMART-goals that they evaluated and revised.

**Figure 1.**
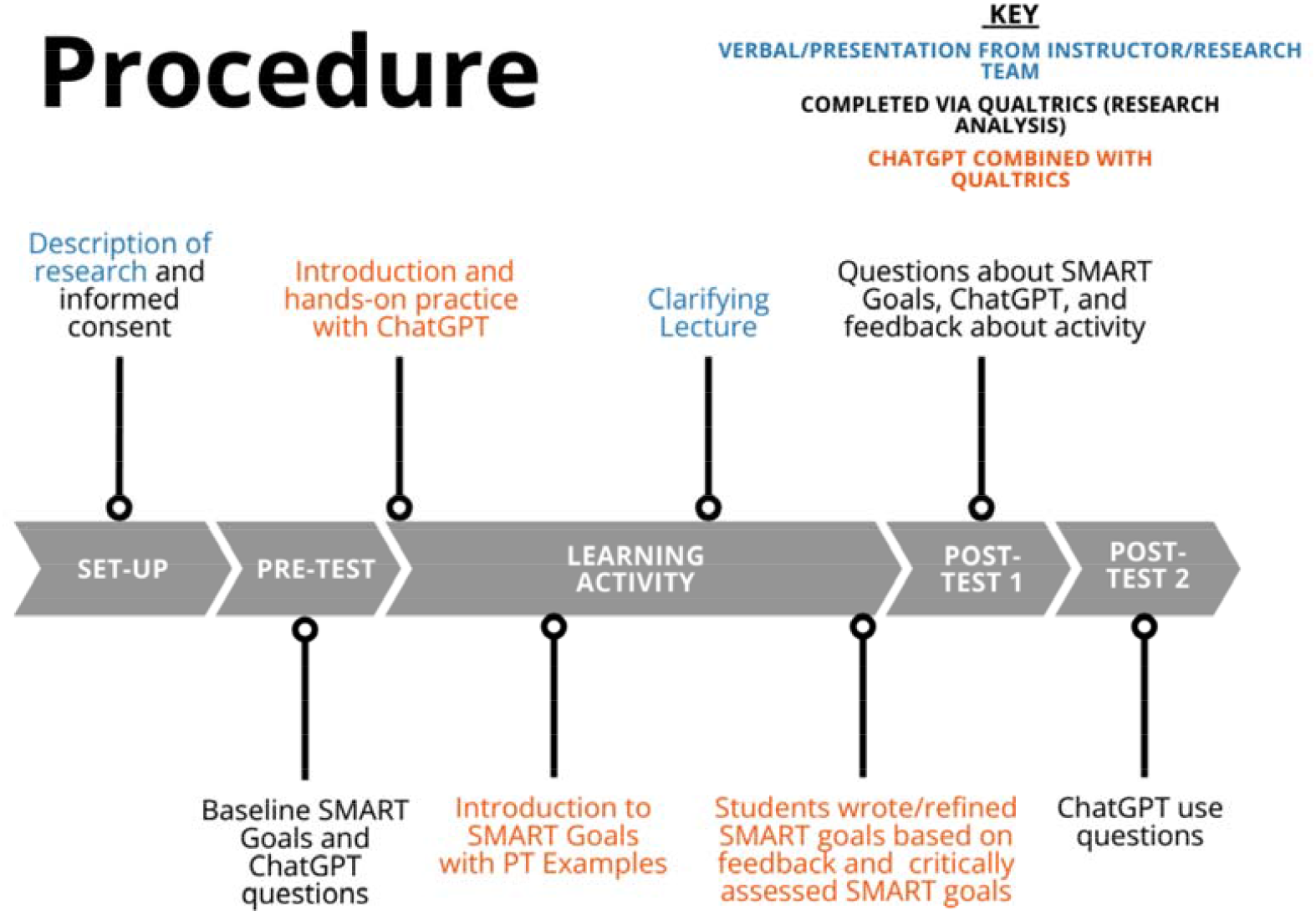
Schematic of learning activity segments and chronology.

### Mixed-Methods Evaluation of the Activity

We administered pre- and post-tests (via Qualtrics, approximately five minutes each) immediately before and after the learning activity. These served to measure the ability of this activity to enhance students’ knowledge of, and confidence in, crafting and revising goals based on clinical scenarios. Additionally, these instruments provided insights into students’ experiences, preferences, and feedback. A second post-test was administered two weeks after the learning experience to gather information regarding students’ ChatGPT usage after the activity. The instruments used for these assessments are available in Appendix 1.

### Statistical Analysis and Data Visualization

For the quantitative data, we calculated descriptive statistics (frequencies and percentages of responses per category) and when appropriate, employed paired t-tests to identify differences between pre-test and post-test responses. We also used emergent coding to elucidate themes from the open-ended survey questions. This coding was conducted by two authors (JES and BLW) and continued until consensus was reached and a final number of responses per category could be determined. We conducted statistical analysis and data visualization in Prism GraphPad (v10).

## Findings

### Knowledge and Skills Related to SMART GOALS

Before the activity, students (n=48) indicated basic understandings of SMART goals and their use in physical therapy (Figure 2). For example, while the majority (87.5%, Figure 2A insert) of students could correctly identify SMART as being an acronym for “specific”, “measurable”, “attainable”, “relevant”, and “time-bound”, they could only, on average, identify about half of the reasons why PTs utilize SMART goals in practice (Figure 2D). Additionally, while most students could recognize a well-written SMART goal, they were not confident in their own abilities to write SMART goals based on a patient scenario (Figure 2C).

**Figure 2.**
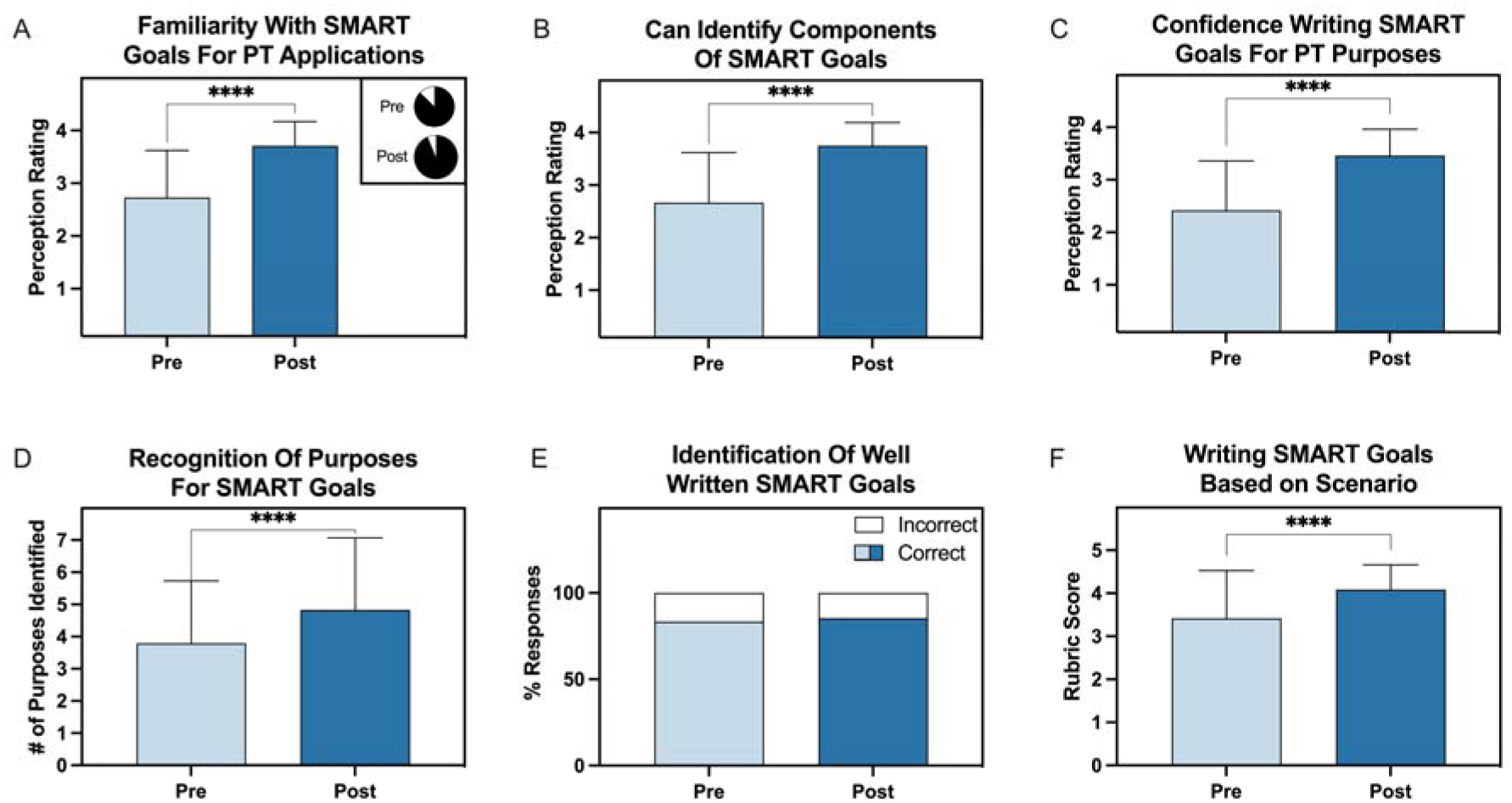
Comparisons between pre-test and post-test responses indicate ways in which the activity contributed to learning and confidence gains. A-C) Students rated their agreement or disagreement with these 3 statements on a 4-point scale (1 = strong disagreement, 2 = slight disagreement, 3 = slight agreement, 4 = strong agreement): “I am familiar with the term SMART goals as it pertains to PT”, “I can identify the components of SMART goals”, and “I feel confident that I can write SMART goals for PT applications.” The pie chart insert in A shows the percentage of students who correctly identified the terms in the SMART acronym correctly (black = correct, white = incorrect). D) Student responses to a question that asked, “Why do Physical Therapists write SMART goals” with 7 options that were all correct. E) Percentage of students who correctly (blue, light blue = pre-test, dark blue = post-test) identified the well-written SMART goal from a multiple-choice question. F) Students were presented with a patient scenario and asked to write a SMART goal. Their submissions were evaluated based on a rubric (Appendix 1) that was also used for the final exam of the course. **** p<0.00001 based on one-tailed paired t-tests. Data shown as mean with bars representing standard deviation.

Responses to the post-test indicate that the activity contributed significantly to students developing increased knowledge and skills in writing and applying the SMART goal framework (Figure 2, p<0.0001) in all areas except identifying well-written SMART goals, the latter being relatively high even before the activity (Figure 2E). After the activity, students also indicated increased confidence regarding their own knowledge and skills (Figure 2 A-C). Compared to the pre-test, responses on the post-test also indicated that more students were able to correctly identify the components of the SMART goal acronym (93.8%, Figure 2A inserts) and ways in which SMART goals are used in PT (m=4.8 purposes, Figure 2D). Student-written SMART goals were scored higher according to the grading rubric (Appendix 1) after the learning activity and their questions for the instructor centered around high-ordered skills such as how to know if the SMART goals are attainable, how to write them concisely, and whether to create separate goals for each of a patient’s impairments/activity limitations (data not shown).

### Perceptions of ChatGPT

The majority of students (56.3%) had never used ChatGPT before the activity and only one student indicated that they had frequently used this tool (Figure 3A). Students were most familiar with concepts of writing prompts, regenerating responses, and asking follow-up questions in ChatGPT, but many were not aware of the ability to generate and share direct URL links to the prompts and AI-responses. Before the activity, students identified a variety of uses for ChatGPT in PT as a clinical specialty and for PT students that included supporting clinical tasks, considering alternative ideas, knowledge expansion, and creating study materials (Table 1 in Appendix 3).

**Figure 3.**
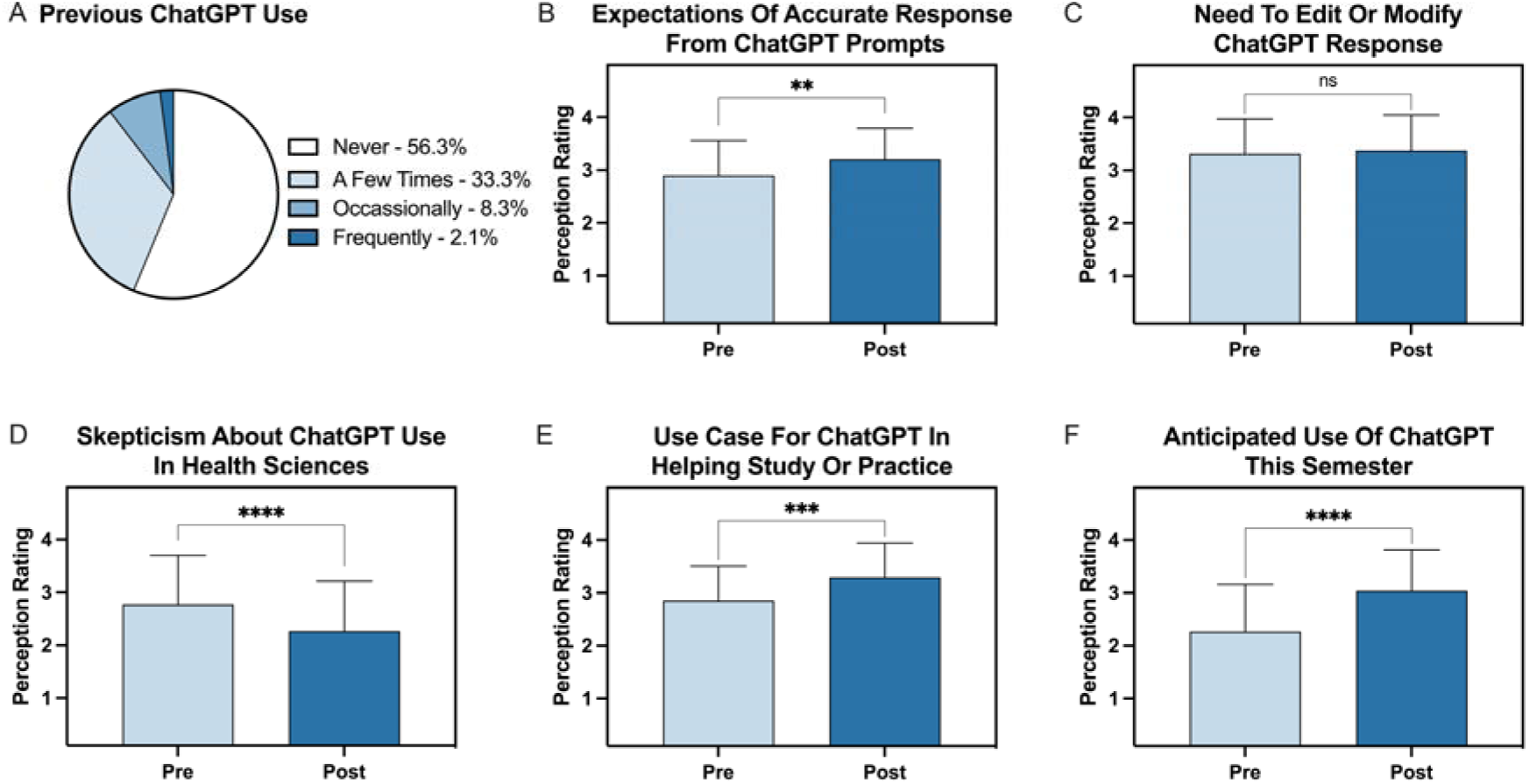
Student responses regarding their use and perceptions of ChatGPT before and after the activity. A) Student responses indicating their frequency of ChatGPT use prior to this activity. B-F) Students rated their agreement or disagreement with these statements on a 4-point scale (1 = strong disagreement, 2 = slight disagreement, 3 = slight agreement, 4 = strong agreement): “I expect that ChatGPT will give me an accurate answer for the questions that I pose to it.”, “If I ask ChatGPT to generate a response, I anticipate I would need to modify or edit the response in some way before using it.”, “I am skeptical about using ChatGPT for applications related to health sciences.”, “I could see a use case for ChatGPT in helping me study or practice skills throughout my degree.” and “Outside of this activity, I anticipate using ChatGPT for personal or educational uses between now and the end of the semester.” n.s. p >0.05, ** p<0.01, *** p<0.001, **** p<0.0001 based on two-tailed paired t-tests. Data shown as mean with bars representing standard deviation.

Comparing results from the pre- and post-tests, it can be seen that using ChatGPT increased students’ expectations of accurate answers from the LLM (p<0.01, Figure 3B) and also increased their ability to see value and practical applications for using ChatGPT to assist with studying or practicing skills both long-term and short-term during their training (p<0.001, Figure 3E-F). To this end, students’ skepticism about the use of ChatGPT in health sciences decreased as a function of the in-class activity (p<0.0001, Figure 3D). However, students’ recognition of the need to edit or modify responses outputted by ChatGPT remained unchanged (p>0.5, Figure 3C).

### Experiences With the Learning Activity

When asked about their experiences with the activity, students indicated slight to strong agreement (Likert values 3 and 4 respectively, Figure 4A) that the activity was helpful (m=3.7), engaging (m=3.5), motivating (m=3.2), enjoyable (m=3.5), and useful (m=3.6). In particular, students found the ability to use ChatGPT as a virtual tutor during the activity to be most helpful (Figure 4B, Table 2 in Appendix 3).

**Figure 4.**
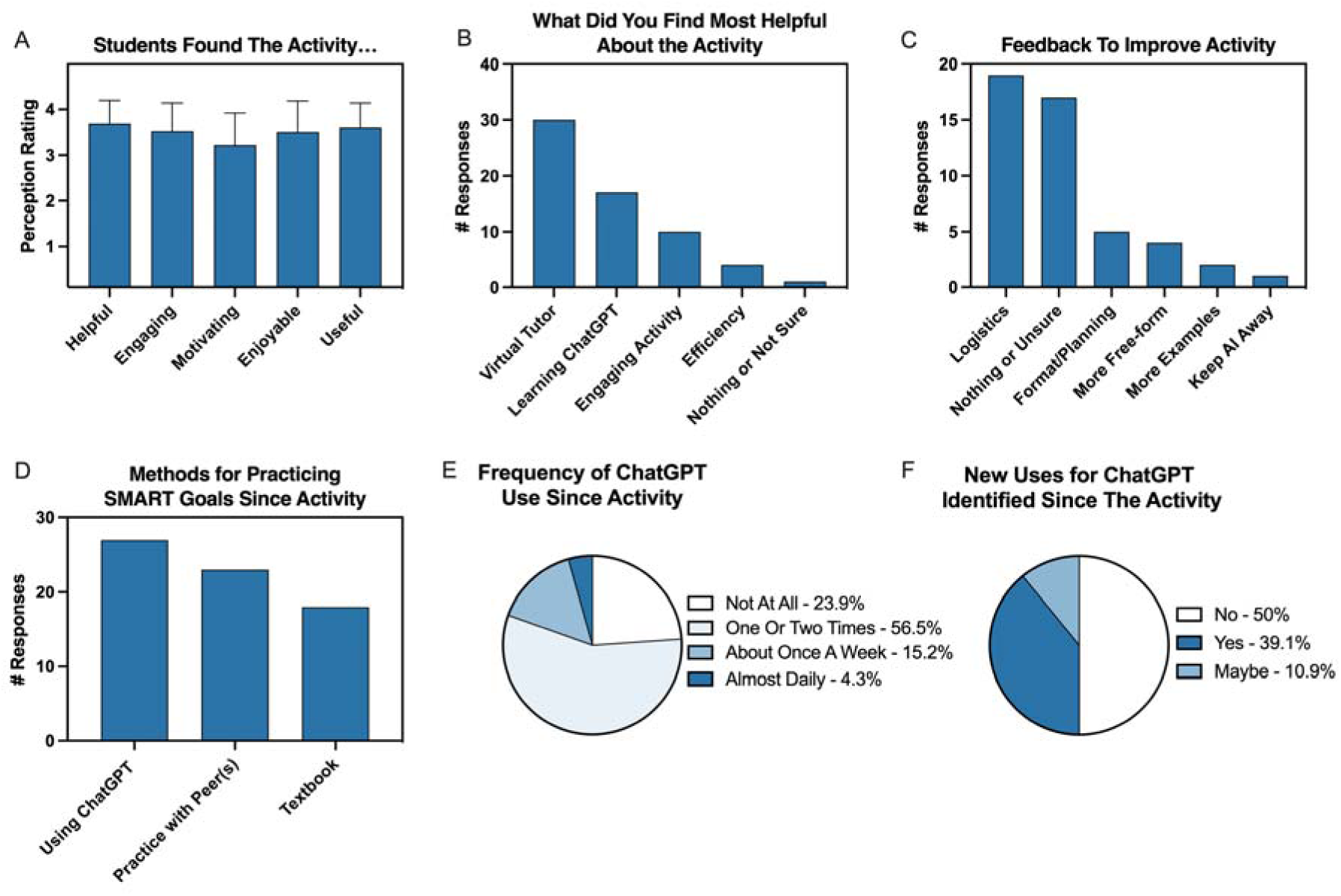
Student responses regarding their perceptions of the activity and their post-activity behaviors. A) Student responses indicated agreement or disagreement the statements, “I found this activity helpful in learning about SMART goals”, “I found this activity engaging”, “Using ChatGPT helped motivate me in this activity.”, “I did not like this activity [reverse scored]”, and “The feedback from ChatGPT helped me to refine the SMART goals that I wrote”. B) Number of student responses in each thematic category related to the elements of the activity they found most helpful. C) Number of student responses in each thematic category related to suggestions for improvements to the activity. D) Number of student responses to the question, “Since the activity in class related to learning about SMART goals, how have you practiced those skills or continued to learn about goal writing in PT?”. E) Frequency of ChatGPT use in the two weeks after the in-class activity. F) Percentage of student responses to a question asking if students have found new uses for ChatGPT since the activity.

Students also enjoyed learning more about ChatGPT and expressed that this was an engaging activity. In contrast, students’ dislikes about the activity (Table 3 in Appendix 3) encompassed issues related to ChatGPT, such as misalignment with students’ preferences, as well as logistical challenges. Specifically, a technical problem emerged when a large number of students attempted to create ChatGPT accounts simultaneously in one location. Additionally, some students found the activity was not challenging enough. While 17 students did not indicate their recommendations for improvements to the activities, the remainder of learners offered suggestions for improving the experience (Figure 4C, Table 4 in Appendix 3) that fell into several categories including logistics, format/planning, making the activity more self-paced or free-form, and including additional examples.

Over the two weeks after the in-class activity, 27 students indicated that they continued to use ChatGPT to help them practice writing or modifying SMART goals (Figure 4D). Additionally, students worked with peers or used the textbook to get additional practice. During that same time frame, the majority of students (76%) reported using ChatGPT again (Figure 4E) and 50% found new or potentially new applications for ChatGPT (Figure 4F) that mostly pertained to educational or personal use.

## Discussion

While previous literature has indicated the potential for ChatGPT and other AI tools to support documentation processes in the healthcare space^24–27^, to the best of our knowledge, this represents the first use of a LLM to support learning related to clinical documentation in the classroom. We designed this activity to intentionally leverage ChatGPT to build an interactive and engaging activity where PT students could learn about the SMART goal framework and its use in PT, while also getting hands on practice and real-time feedback writing and modifying SMART goals based on scenarios. Because it was unknown how many students had experience using ChatGPT and what their comfort would be with prompt engineering, we built in opportunities for students to explore the tool before getting started and we provided instructor-designed prompts throughout the activity to ensure students had consistent learning experiences, regardless of their prior use of AI-based tools. The qualitative and quantitative data indicate that this activity served as a valuable educational tool, functioning as a virtual tutor, to support students in comprehending and formulating SMART goals. The evidence suggests an enhancement in students’ confidence levels, as well as positive shifts in their perceptions of their knowledge, skills, and abilities. This impact is likely attributed, in part, to the engaging and motivating nature of the activity and the immediate feedback provided by ChatGPT. In addition to fostering a deeper understanding of SMART goals, the activity facilitated an exploration of ChatGPT’s capabilities and limitations. It emphasized the importance of critically evaluating the chatbot’s responses. Students discovered that this tool is best used during the brainstorming process and that their active inputs and modifications are needed to develop clinically relevant goals written in ways that align to PT standards of practice. The clarifying lecture also provided an opportunity to discuss the differences between how ChatGPT often generated goals (as a bulleted list or paragraph) and how PTs actually write goals (in sentence form). Together, the findings suggest that the activity was able to address the learning challenge and support students in achieving the learning objectives aligned to this lesson.

As much as students indicated that they liked the activity, the feedback from students also illuminated several opportunities for future iterations of this activity. A subset of students revealed their preferences for live class sessions to focus more on direct instruction from the faculty member or to involve peer-peer discussion. These responses highlight two different possible paths for using this activity in the future. First, the activity could be adjusted easily into a homework or pre-class assignment where students interact with ChatGPT outside of class time to learn more about SMART goals and practice writing the goals before coming to class. Alternatively, in the original activity, group work was intentionally minimized to enhance our ability to measure individual learning gains resulting from the activity. In the future, however, the activity could be adjusted to include a significant collaborative component such that students could learn from the perspectives and opinions of their peers while still leveraging ChatGPT in the role of a peer-tutor within the group. Other responses suggested that some students felt that ChatGPT was doing the thinking for them, found the activity to be too closed in nature, or did not present enough of a challenge. For future iterations, adjustments to the activity and its instructions could enhance clarity about the expectation for students to engage with ChatGPT in a “brains on” manner by actively participating in “conversation” with, and critically evaluating, responses from ChatGPT. Moreover, facilitating an open-ended exploration of ChatGPT to deepen understanding of the topic and introducing additional layers of complexity to the segment where students formulate and modify SMART goals might address these concerns. Lastly, many students commented on logistics. We built in time as part of the activity for students to make OpenAI accounts if they didn’t already have one. What we didn’t realize at the time, was that the website can become unresponsive and fail to load when numerous people try to create an account simultaneously. This caused a delay in the activity that students found frustrating. We made several attempts to troubleshoot and reduce the impacts of this technical challenge. Despite our efforts, however, some students started the activity using tablets or laptops with a partner while the website loaded. In the future, this challenge may be avoided by requesting that students create OpenAI accounts prior to arriving in class.

Additional study would be necessary to assess the limitations of this study and to explore the long-term effects of this activity in terms of knowledge retention and its impact on students’ clinical performance. However, based on the data, we believe that this activity positively influenced the learners, encouraging its further use in this instructional setting and for other educational purposes. Further testing could also validate the impact of other LLMs including Gemini, Bing, and the more advanced versions of ChatGPT (ChatGPT 4.0) for their use in this activity^28^. Additionally, we believe that the activity presented here is easy for other educators to implement, and is modifiable and scalable, which makes it easy to transfer to other courses or disciplines. However, while implementing technology, we emphasize the importance of intentionally aligning its use with the objectives and context of the class. It is important to avoid incorporating technology merely for its own sake, but rather, to utilize it deliberately while aiming to create an active learning experience and foster the development of meta-skills, including metacognition and critical thinking, as advocated by the TPACK framework^29^. Furthermore, it’s important to both model and teach students about responsible use of AI-based tools and to help them navigate the opportunities and the risks these tools offer while recognizing that these tools will be transformational in higher education and clinical practice.

## Conclusions

This study represents a novel active learning experience that leveraged ChatGPT as a virtual tutor to support student learning outcomes related to clinical documentation. The data demonstrate that students enjoyed the activity which is scalable and adaptable for other health education topics. The data further reveal student preferences and experiences related to AI-based tools in education and health care.

## Data Availability

The data used in this study are available upon written request to the corresponding author.

## Acknowledgements

The authors would like to thank the students who participated in this activity for their support and generosity in providing their feedback about the activity. Additionally, we would like to thank Drs. Quincy Conley and Lori Bordenave for their support of this research.

## Data Availability Statement

The data used in this study are available upon written request to the corresponding author.

## Author Contributions

**Julie Speer:** Conceptualization; Formal Analysis; Investigation; Methodology; Project Administration; Supervision; Visualization; Writing – Original Draft Preparation; Writing – Review and Editing. **Sara Parker:** Conceptualization; Formal Analysis; Investigation; Methodology; Project Administration; Writing – Original Draft Preparation; Writing – Review and Editing. **Brittany Williams:** Conceptualization; Formal Analysis; Investigation; Methodology; Project Administration; Writing – Original Draft Preparation; Writing – Review and Editing.

## Ethical Approval

No patient-sensitive data were used in this study. The research protocol was approved as an exempt research study by the A.T. Still University Arizona Institutional Review Board (#2023-142) and participants provided consent to have their responses analyzed through a signed informed consent process.

## References

1. Effgen SK, Chiarello L, Milbourne SA. Updated competencies for physical therapists working in schools. Pediatr Phys Ther. 2007;19(4):266–274. doi:10.1097/PEP.0b013e318158ce90

2. Timmerberg JF, Dole R, Silberman N, et al. Physical therapist student readiness for entrance into the first full-time clinical experience: a delphi study. Phys Ther. 2019;99(2):131–146. doi:10.1093/ptj/pzy134

3. Shamus E, Stern DF. Effective Documentation for Physical Therapy Professionals. 2nd ed. McGraw-Hill Medical; 2011. https://search.ebscohost.com/login.aspx?direct=true&AuthType=shib&db=cat02627a&AN=atsmo.b3393819&site=eds-live&custid=s4165981

4. Ho Y-X, Gadd CS, Kohorst KL, Rosenbloom ST. A qualitative analysis evaluating the purposes and practices of clinical documentation. Appl Clin Inform. 2014;5(1):153–168. doi:10.4338/ACI-2013-10-RA-0081

5. Physical therapy (residential), DPT. https://catalog.atsu.edu/preview_program.php?catoid=23&poid=510#courses

6. Farzandipour M, Meidani Z, Rangraz Jeddi F, et al. A pilot study of the impact of an educational intervention aimed at improving medical record documentation. J R Coll Physicians Edinb. 2013;43(1):29–34. doi:10.4997/JRCPE.2013.106

7. Dutton LL, Sellheim DO. The Informal and Hidden Curriculum in Physical Therapist Education. J Phys Ther Educ. 2014;28(3):50–63. doi:10.1097/00001416-201407000-00008

8. Garcia A, Revere L, Sharath S, Kougias P. Implications of clinical documentation (in)accuracy: a pilot study among general surgery residents. Hosp Top. 2017;95(2):27–31. doi:10.1080/00185868.2017.1300471

9. Ridyard E, Street E. Evaluating the quality of medical documentation at a university teaching hospital. BMJ Qual Improv Reports. 2015;4(1):u208052.w3253. doi:10.1136/bmjquality.u208052.w3253

10. Benoit JRA. ChatGPT for clinical vignette generation, revision, and evaluation. medRxiv. Published online 2023. https://www.medrxiv.org/content/10.1101/2023.02.04.23285478v1%0Ahttps://www.medrxiv.org/content/10.1101/2023.02.04.23285478v1.abstract [preprint]

11. Dai W, Lin J, Jin H, et al. Can large language models provide feedback to students? A case study on ChatGPT. In: 2023 IEEE International Conference on Advanced Learning Technologies (ICALT). IEEE; 2023:323-325. doi:10.1109/ICALT58122.2023.00100

12. Elkassem AA, Smith AD. Potential use cases for ChatGPT in radiology reporting. Am J Roentgenol. 2023;221(3):373–376. doi:10.2214/AJR.23.29198

13. Johnson D, Goodman R, Patrinely J, et al. Assessing the accuracy and reliability of AI-generated medical responses: an evaluation of the Chat-GPT model. Published online 2023. doi:10.21203/rs.3.rs-2566942/v1

14. Lee H. The rise of ChatGPT: exploring its potential in medical education. Anat Sci Educ. 2023;(March):1–6. doi:10.1002/ase.2270

15. Paranjape K, Schinkel M, Nannan Panday R, Car J, Nanayakkara P. Introducing artificial intelligence training in medical education. JMIR Med Educ. 2019;5(2):e16048. doi:10.2196/16048

16. Patel SB, Lam K. ChatGPT: the future of discharge summaries? Lancet Digit Heal. 2023;5(3):e107–e108. doi:10.1016/S2589-7500(23)00021-3

17. Sallam M. ChatGPT utility in healthcare education, research, and practice: systematic review on the promising perspectives and valid concerns. Healthcare. 2023;11(6):887. doi:10.3390/healthcare11060887

18. Zuccon G, Koopman B. Dr ChatGPT, tell me what I want to hear: how prompt knowledge impacts health answer correctness. arXiv. 2023;1(1). doi:10.48550/arXiv.2302.13793[preprint]

19. Liu S, Wright AP, Patterson BL, et al. Using AI-generated suggestions from ChatGPT to optimize clinical decision support. J Am Med Informatics Assoc. 2023;30(7):1237–1245. doi:10.1093/jamia/ocad072

20. Fazlollahi AM, Bakhaidar M, Alsayegh A, et al. Effect of artificial intelligence tutoring vs expert instruction on learning simulated surgical skills among medical students. JAMA Netw Open. 2022;5(2):e2149008. doi:10.1001/jamanetworkopen.2021.49008

21. Huang J, Tan M. The role of ChatGPT in scientific communication: writing better scientific review articles. Am J Cancer Res. 2023;13(4):1148–1154. Accessed February 22, 2024. https://www.ajcr.us/

22. Dennick R. Constructivism: reflections on twenty five years teaching the constructivist approach in medical education. Int J Med Educ. 2016;7:200–205. doi:10.5116/ijme.5763.de11

23. Keller JM. Development and use of the ARCS model of instructional design. J Instr Dev. 1987;10(3):2–10. doi:10.1007/BF02905780

24. Sun GH, Hoelscher SH. The ChatGPT storm and what faculty can do. Nurse Educ. 2023;48(3):119–124. doi:10.1097/NNE.0000000000001390

25. Iftikhar L, Iftikhar MF, Hanif MI. DocGPT: impact of ChatGPT-3 on health services as a virtual doctor. EC Paediatr. 2023;12(3):45–55. https://ecronicon.org/assets/ecpe/pdf/ECPE-12-01277.pdf

26. Clough RAJ, Sparkes WA, Clough OT, Sykes JT, Steventon AT, King K. Transforming healthcare documentation: harnessing the potential of AI to generate discharge summaries. BJGP Open. Published online September 12, 2023:BJGPO.2023.0116. doi:10.3399/BJGPO.2023.0116

27. Decker H, Trang K, Ramirez J, et al. Large language model-based chatbot vs surgeon-generated informed consent documentation for common procedures. JAMA Netw Open. 2023;6(10):e2336997. doi:10.1001/jamanetworkopen.2023.36997

28. Seth I, Lim B, Xie Y, et al. Comparing the Efficacy of Large Language Models ChatGPT, BARD, and Bing AI in Providing Information on Rhinoplasty: An Observational Study. Aesthetic Surg J Open Forum. 2023;5(0):1–9. doi:10.1093/asjof/ojad084

29. Koehler MJ, Mishra P, Cain W. What is technological pedagogical content knowledge (TPACK)? J Educ. 2013;193(3):13–19.

