## Appendix 1 for "Interactive Learning with ChatGPT: Hands-On Practice and Real-Time Feedback in Health Sciences Education for SMART Goal Writing"

Instruments and Activities

### Pre-Test

**Instructions for the pre-test:** It is ok if you don’t know the answer to every question, the learning activity you will engage with today will address these themes. Please respond to the questions below to the best of your knowledge.

Please indicate your agreement or disagreement with the following statements (1 = Strong disagreement, 2 = Slight disagreement, 3 = Slight agreement, 4 = Strong agreement):

|  | 1 - Strong disagreement | 2 - Slight disagreement | 3 - Slight agreement | 4 - Strong agreement |
| --- | --- | --- | --- | --- |
| I am familiar with the term SMART goals as it pertains to PT |  |  |  |  |
| I can identify the components of SMART goals |  |  |  |  |
| I feel confident that I can write SMART goals for PT applications |  |  |  |  |

What does the acronym SMART in SMART Goals stand for?

- Structured, Managed, Appropriate, Responsive, Timed
- Sequential, Motivational, Achievable, Realistic, Time-bound
- Specific, Meaningful, Action-Oriented, Relevant, Timely
- Specific, Measurable, Attainable, Relevant, Time-bound

Why do Physical Therapists write SMART Goals? [Select all that apply]

- For record keeping
- To allow other healthcare providers to see or know what the patient is working on
- To communicate expectations with patients
- To remind the PT what to work on with the patient in the next session
- To help with treatment planning and tracking progress
- To help motivate patients
- For regulatory purposes

Which of the following is a well written SMART goal?

- The patient will walk two miles on a flat sidewalk without assistive device and with no pain or hip instability in three months.
- The patient will regain muscle strength with VMO isometric contraction and straight leg raises to improve gait control.
- The patient will improve standing balance (eyes open and eyes closed) in four weeks.
- The patient will sit unsupported in short-leg sitting to enable upright activities

What suggestions would you make to modify this goal "The patient will demonstrate increased hip flexion during lifting activities" to better comply with the SMART framework?

________________________________________________________________

________________________________________________________________

________________________________________________________________

________________________________________________________________

________________________________________________________________

Based on this truncated patient scenario below, please write a SMART goal.

A 45-year-old male patient named John presents to the physical therapy clinic with complaints of right shoulder pain at 5/10 that has been bothering him for the past four weeks. He reports that the pain initially started gradually and has been worsening over time. John states that he is unsure of any specific incident that may have caused the pain. He describes the pain as a sharp, stabbing sensation that is aggravated with overhead activities and reaching behind his back. He also mentions occasional night pain and difficulty with sleeping on his right side. John denies any numbness, tingling, or weakness in the affected arm. He works as a computer programmer and spends long hours sitting at his desk.

**Range of Motion (ROM):**
Active shoulder flexion: 120 degrees (normal: 180 degrees)
Active shoulder abduction: 100 degrees (normal: 180 degrees)

**Strength Testing (normal is 5/5 for all movements)**
Shoulder abduction: 4/5 (compared to the unaffected side)

________________________________________________________________

________________________________________________________________

________________________________________________________________

________________________________________________________________

________________________________________________________________

Have you used ChatGPT before today for any purpose?

- Never
- A few times, but not often
- Occasionally – a few times a month
- Frequently

Which features of ChatGPT are you familiar with? [Select all that apply]

- Writing prompts
- Sharing chats
- Regenerating responses
- Asking follow-up questions to the AI

How do you think ChatGPT can be useful for PTs or PT students?

________________________________________________________________

________________________________________________________________

Please indicate your agreement or disagreement with the following statements (1 = Strong disagreement, 2 = Slight disagreement, 3 = Slight agreement, 4 = Strong agreement):

|  | 1 - Strong disagreement | 2 - Slight disagreement | 3 - Slight agreement | 4 - Strong agreement |
| --- | --- | --- | --- | --- |
| I expect that ChatGPT will give me an accurate answer for the questions that I pose to it. |  |  |  |  |
| If I ask ChatGPT to generate a response, I anticipate I would need to modify or edit the response in some way before using it. |  |  |  |  |
| I am skeptical about using ChatGPT for applications related to health sciences. |  |  |  |  |
| I could see a use case for ChatGPT in helping me study or practice skills throughout my degree. |  |  |  |  |
| Outside of this activity, I anticipate using ChatGPT for personal or educational uses between now and the end of the semester. |  |  |  |  |

### ChatGPT Playground

**Instructions:** Please follow the instructions to start using ChatGPT and testing out some of its key features that we will use in today’s activity.

**Step 1:** Before we get started, please either log in or create an account with ChatGPT at https://chat.openai.com (please open in a separate tab or window). 
  

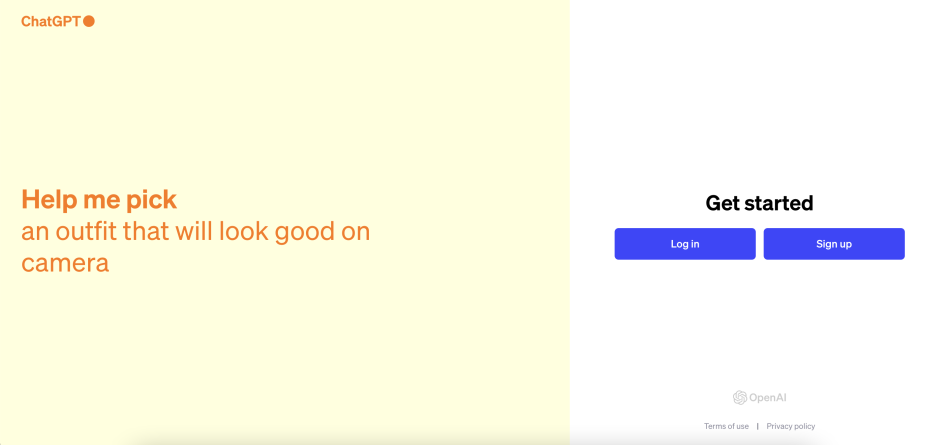

 Image screenshot from ChatGPT. Retrieved from: https://chat.openai.com on August 17, 2023.

If you are creating a new account, please note that you will need to access your email to click on the verification email.


You will also be prompted to supply your birthday and your phone number (used to further verify your account). You will not be asked to share any of this information as part of the research study. Once you log in for the first time, a quick tutorial will appear. Click next to move through the overview.

**Step 2:** From the home screen, type or copy/paste the following prompt into ChatGPT: “What is the state flower of Arizona?”

           
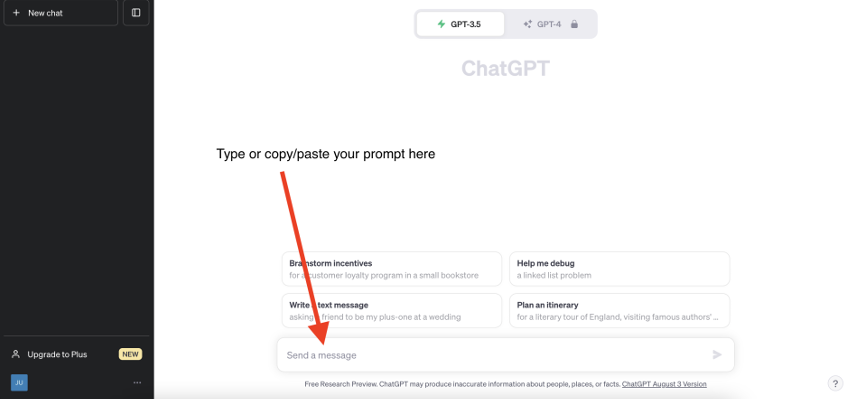

 Image screenshot from ChatGPT. Retrieved from: https://chat.openai.com on August 17, 2023.

**Step 3:** Allow the response from ChatGPT to generate and read the response.

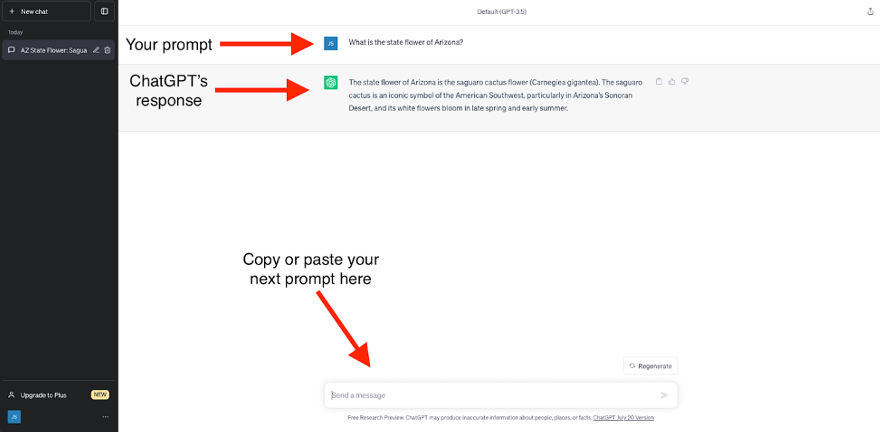

 Image screenshot from ChatGPT. Retrieved from: https://chat.openai.com on August 16, 2023.

**Step 4:** Repeat the previous step (step 3) with the following prompts (which are bolded):

 **- Tell me more about the state flower of Arizona.**

 Why did we ask you to do this? What does this show? As you can see from this prompt and response, you can ask ChatGPT for follow up questions about what it has previously told you in the chat.

 - **Rewrite that response using only 100 words.** 

 Why did we ask you to do this? What does this show? This example shows that we can ask ChatGPT to do follow up steps and to summarize information with constraints like word count or with information about who the audience is intended for.

 - **Let me check if I understand. Is this correct? "The state flower of Arizona is the Saguaro Cactus. The small flowers bloom in the springtime and last about 1 day only. It was chosen as the state flower because it represents the resilience and beauty of the American Northeast."**

 Why did we ask you to do this? What does this show? We can ask ChatGPT for feedback about responses we write. So here we can ask it to check whether our answer has captured the information it has previously told us about.

 - **Where do you recommend that I learn more about the Saguaro Cactus Blossom?**

 Why did we ask you to do this? What does this show? You can ask ChatGPT for recommendations about future reading or additional materials.

 - **Ask me a question to test my understanding of the state flower of Arizona.** 

Then type in your response to the question ChatGPT generates for you and ask if your response is correct. Please do this all in one prompt like shown below.

 Why did we ask you to do this? What does this show? You can ask ChatGPT to generate questions to help you summarize information or check your own understanding.

  
 

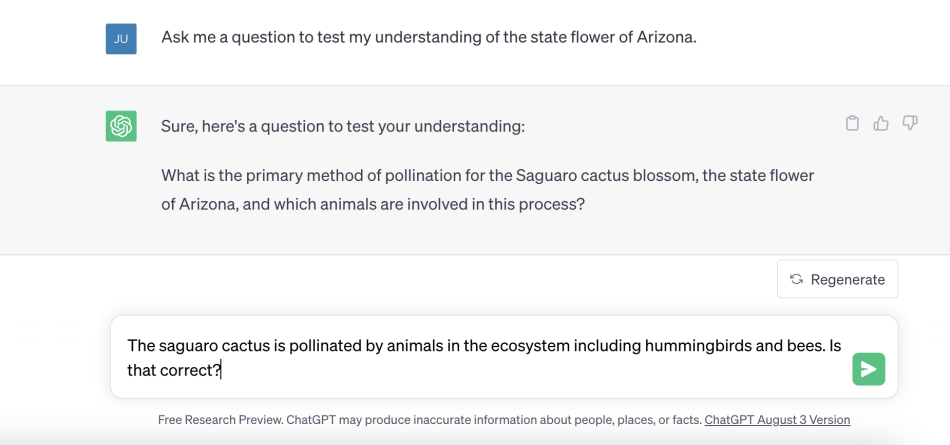

  Image screenshot from ChatGPT. Retrieved from: https://chat.openai.com on August 17, 2023.

How did ChatGPT do in rating your response?

- ChatGPT labeled my response as being correct, but it was incorrect.
- ChatGPT labeled my response as being correct and it was correct.
- ChatGPT labeled my response as incorrect but it was correct.
- ChatGPT labeled my response as incorrect and it was incorrect.

**Step 5:** As shown in the GIF below, please rename this chat as “ChatGPT Playground” and share the URL to your chat in the text box below.

Click for written directions: Step 1. Scroll to the top of your chat. Step 2. From the left-hand panel, find your current chat and click on the pencil icon. Step 3. Delete the text that is currently there and replace it with, 'ChatGPT Playground'. Step 4. Click the check icon to save the new name. Step 5. In the top right-hand corner of the screen find the share icon (box with an arrow). Click on the icon. Step 6. Click the green Copy Link button and paste the URL in the text box below.

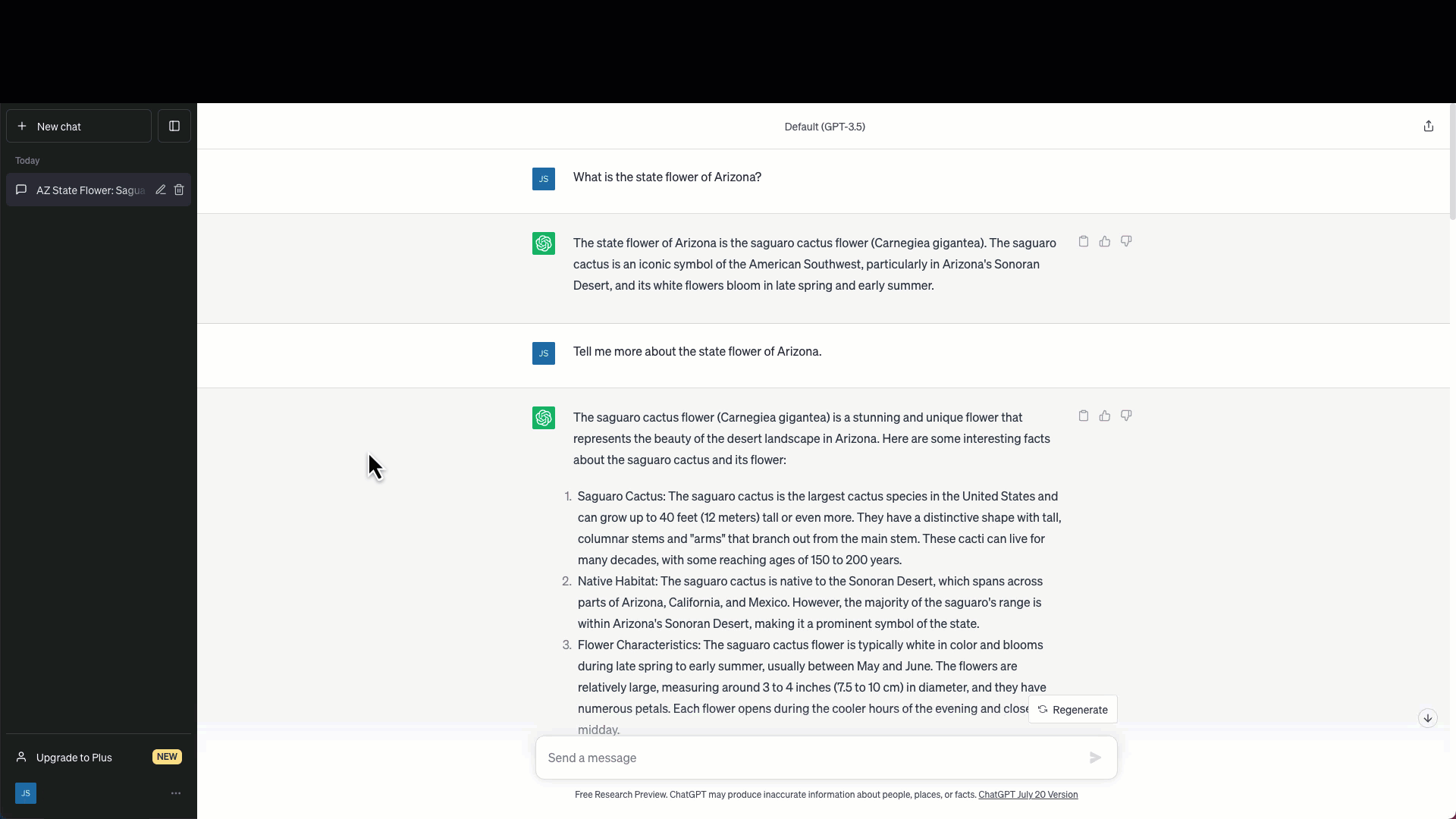

 Screen capture showing ChatGPT. Retrieved from: https://chat.openai.com on August 1, 2023.

The URL for my ChatGPT Playground chat is:

________________________________________________________________

**Step 6:** To prepare for the next step in the activity, please create a new chat by clicking the “+ New Chat” button at the top left of the screen.

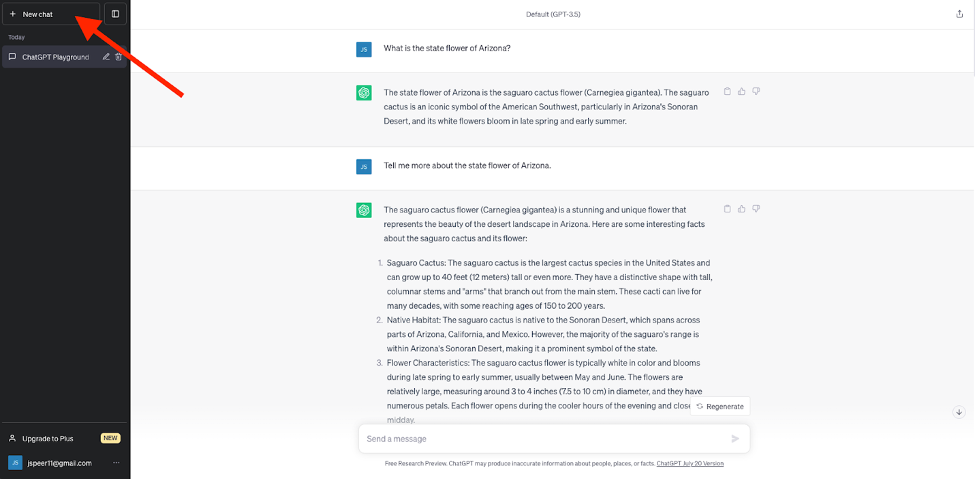

 Screen capture showing ChatGPT. Retrieved from: https://chat.openai.com on August 16, 2023.

### Learning Activity 1

**Instructions:** Please follow the steps to use ChatGPT to learn about SMART goals and their use in physical therapy.

- Your first and last name__________________________________________________

**Step 1:** In your new chat, please type (or copy/paste) in the following prompts sequentially. Please read the text that ChatGPT generates thoroughly before asking the next prompt.

●	What are SMART goals?

●	Can you distill that answer down to about 200 words?

●	Why are SMART goals a common framework in the health sciences as part of

documentation?

●	Are SMART goals the only framework for goal setting in the health sciences?

●	Can you give me an analogy to help me understand SMART goals in healthcare?

●	How do I write a SMART goal in physical therapy?

●	Can you give me some prompts that will help me write SMART goals for physical

therapy applications?

●	Can you give me an example of a SMART goal for a physical therapy application?

●	Ok so let's say you have a patient who has come to the physical therapist with acute

pain in their shoulder following a rotator cuff tear. What might be a SMART goal that

would relate to that example?

**Step 2:** Following the steps we performed before, please rename this chat, “Learning Activity Part 1” and share the link to your chat here:

________________________________________________________________

### Learning Activity 2

**Instructions:** Please follow the steps outlined below to use ChatGPT to practice writing and revising SMART goals.

- Your name (first and last name) __________________________________________________

**Part 1:** In a new chat, please type in the following prompts *sequentially*. Please follow the directions between each prompt before moving on to the next.

 1. Enter this *1st prompt* into ChatGPT: “Create a new patient for physical therapy with subjective findings and specific objective findings of ROM, strength and special tests, palpation; no diagnosis or summary but include normal values. Can you include specific measurements of ROM and pain in 100 words or less?” Based on that scenario generated by ChatGPT, please write a potential SMART goal for that patient.

2. Check your SMART goal with ChatGPT by typing this *2nd prompt*: “Does this goal capture all components of SMART goals? "*Insert Your Goal Here*". 

3. Ask ChatGPT a *3rd prompt*: “Are there modifications that should be made to improve this SMART goal?”

4. Based on your feedback from ChatGPT and your own knowledge of SMART goals, what is your final SMART goal for this patient:

________________________________________________________________

________________________________________________________________

________________________________________________________________

________________________________________________________________

________________________________________________________________

**Part 2.** In the same chat, please type in the following prompts sequentially:

1. 1st Prompt: "In 100 words, please give me a new patient scenario about a patient who needs a physical therapist."

2. Once created, ask ChatGPT to write a SMART goal for that patient by using the following 2nd prompt: “Write a SMART goal for that patient”

3. Do you agree with ChatGPT's SMART goal for the patient?

________________________________________________________________

________________________________________________________________

________________________________________________________________

________________________________________________________________

________________________________________________________________

4. Would you make any modifications to ChatGPT's SMART goal for this patient?

________________________________________________________________

________________________________________________________________

________________________________________________________________

________________________________________________________________

________________________________________________________________

5. What would your final SMART goal for this patient be?

________________________________________________________________

________________________________________________________________

**Part 3:** In the same chat, please type in the following prompts sequentially.

1. 1st prompt: "In 100 words, please give me a new patient scenario about a patient who needs a physical therapist."

2. Ask ChatGPT to write a SMART goal that is missing one or more components but make sure ChatGPT doesn’t tell you what is missing using this 2nd prompt: “Please write a SMART goal for this patient that is missing one or more components of the SMART goal framework. Do not tell me what is missing or wrong and do not correct the goal.”

3. What components are incorrect or missing?

________________________________________________________________

________________________________________________________________

________________________________________________________________

________________________________________________________________

________________________________________________________________

4. How would you fix this SMART goal? Rewrite the goal:

________________________________________________________________

________________________________________________________________

________________________________________________________________

________________________________________________________________

________________________________________________________________

5. Then ask ChatGPT to help you check what you drafted by using the 3rd prompt: “Does this goal better capture all of the SMART goal components for this patient, "*Insert Your Goal Here.*"

6. Further check your SMART goal by typing in 4th prompt: “Would you rewrite the SMART goal to better capture the SMART goal framework?”

7. Based on the feedback you have gotten from ChatGPT, what is your final SMART goal for this patient scenario:

________________________________________________________________

________________________________________________________________

________________________________________________________________

________________________________________________________________

**Part 4:** *If time allows, please complete this activity*. *There will be no penalty if not completed.*

1. Write your own patient scenario in about 50-100 words.

2. Based on that scenario, what SMART goal would you write for that patient:

________________________________________________________________

________________________________________________________________

________________________________________________________________

________________________________________________________________

________________________________________________________________

3. Check that goal with ChatGPT by using the following 1st prompt, “Here is a scenario about a patient: “*Insert your scenario here.*” Does this SMART goal include all the necessary components? “*Insert your goal here*”. ***This is all included as one prompt.**

4. If ChatGPT suggests your goal has included all of the components, please ask it for the following 2nd prompt: “Would you recommend any edits to improve this SMART goal?”

5. Based on that feedback, what is your final SMART goal for your scenario? If ChatGPT suggests your SMART goal was missing components, with the feedback it provided, what is your final SMART goal for this patient?

________________________________________________________________

________________________________________________________________

________________________________________________________________

________________________________________________________________

________________________________________________________________

6. Following the steps we performed before, please rename this chat, “Learning Activity Part 2” and share the link to your chat here:

________________________________________________________________

### Post-Test 1

**Instructions:** Please answer the questions below to the best of your ability and as honestly as possible.

- Your name (first and last name) __________________________________________________
- Did you work with a partner on the ChatGPT activity? If so, please indicate their names here __________________________________________________

Please indicate your agreement or disagreement with the following statements (1 = Strong disagreement, 2 = Slight disagreement, 3 = Slight agreement, 4 = Strong agreement):

|  | 1 - Strong disagreement | 2 - Slight disagreement | 3 - Slight agreement | 4 - Strong agreement |
| --- | --- | --- | --- | --- |
| I am familiar with the term SMART goals as it pertains to PT |  |  |  |  |
| I can identify the components of SMART goals |  |  |  |  |
| I feel confident that I can write SMART goals for PT applications |  |  |  |  |

What does the acronym SMART in SMART Goals stand for?

- Structured, Managed, Appropriate, Responsive, Timed
- Specific, Measurable, Attainable, Relevant, Time-bound
- Sequential, Motivational, Achievable, Realistic, Time-bound
- Specific, Meaningful, Action-Oriented, Relevant, Timely

Why do Physical Therapists write SMART Goals? [Select all that apply]

- For record keeping
- To allow other healthcare providers to see or know what the patient is working on
- To communicate expectations with patients
- To remind the PT what to work on with the patient in the next session
- To help with treatment planning and tracking progress
- To help motivate patients
- For regulatory purposes

Which of the following is the best SMART goal?

- Pt will improve standing balance in 2 weeks.
- Pt will perform sit to/from stand with minA for 4/5 trials in within 2 wks.
- Pt will progress from walker to cane in clinic within 3 weeks.
- Pt will not have pain when reaching into cupboard.

What suggestions would you make to modify this goal to better comply with the SMART framework? “Pt will ascend and descend stairs within 2 weeks.”

________________________________________________________________

________________________________________________________________

________________________________________________________________

________________________________________________________________

________________________________________________________________

Based on this truncated patient scenario below, please write a SMART goal:

Pt is 35 y/o male who reports dull, achy pain in his right shoulder for 3 weeks, aggravated by lifting. Pain intensity is 6/10, interfering with work and sleep. He works as an office manager.

**Range of Motion (ROM):**
Right shoulder flexion: 140° (normal: 180°)
Abduction: 110° (normal: 180°)
External rotation: 45° (normal: 90°)

**Strength:**
Right supraspinatus (Jobe's Test): Weakness noted at 4/5 (normal: 5/5).

________________________________________________________________

________________________________________________________________

________________________________________________________________

________________________________________________________________

Has this activity given you any new ideas about how you can use ChatGPT related to your studies or PT applications?

- Yes
- Maybe
- No

Please indicate your agreement or disagreement with the following statements (1 = Strong disagreement, 2 = Slight disagreement, 3 = Slight agreement, 4 = Strong agreement):

|  | 1 - Strong disagreement | 2 - Slight disagreement | 3 - Slight agreement | 4 - Strong agreement |
| --- | --- | --- | --- | --- |
| I expect that ChatGPT will give me an accurate answer for the questions that I pose to it. |  |  |  |  |
| If I ask ChatGPT to generate a response, I anticipate I would need to modify or edit the response in some way before using it. |  |  |  |  |
| I am skeptical about using ChatGPT for applications related to health sciences. |  |  |  |  |
| I could see a use case for ChatGPT in helping me study or practice skills throughout my degree. |  |  |  |  |
| Outside of this activity, I anticipate using ChatGPT for personal or educational uses between now and the end of the semester. |  |  |  |  |

Based on the activity you just completed, please indicate your agreement or disagreement with the following statements (1 = Strong disagreement, 2 = Slight disagreement, 3 = Slight agreement, 4 = Strong agreement):

|  | 1 - Strong disagreement | 2 - Slight disagreement | 3 - Slight agreement | 4 - Strong agreement |
| --- | --- | --- | --- | --- |
| I found this activity helpful in learning about SMART goals |  |  |  |  |
| I found this activity engaging |  |  |  |  |
| Using ChatGPT helped motivate me in this activity |  |  |  |  |
| I did not like this activity |  |  |  |  |
| The feedback from ChatGPT helped me to refine the SMART goals that I wrote |  |  |  |  |

What did you find least helpful about this activity?

________________________________________________________________

________________________________________________________________

________________________________________________________________

What did you find most helpful about this activity?

________________________________________________________________

________________________________________________________________

________________________________________________________________

What questions do you still have about SMART goals?

________________________________________________________________

________________________________________________________________

________________________________________________________________

________________________________________________________________

________________________________________________________________

Do you have any suggestions about how to improve this activity for future students?

________________________________________________________________

________________________________________________________________

________________________________________________________________

________________________________________________________________

________________________________________________________________

### Post-Test 2

**Instructions:** Please answer the following questions as honestly as possible.

- Your name (first and last name) __________________________________________________

Since the activity in class related to learning about SMART goals, how have you practiced those skills or continued to learn about goal writing in PT? [select all that apply]

- Practicing with examples from the textbook
- Practicing with a peer or in a study group
- Attending office hours or talking with the instructor about SMART goals
- Using ChatGPT to practice on my own

Since the ChatGPT activity we did in class together, how often have you used ChatGPT for any purpose?

- Not at all
- One or two times
- About once a week
- Almost daily

Since the ChatGPT activity we did together in class, have you identified any new applications for ChatGPT? If yes or maybe, feel free to explain in the text boxes for those options respectively.

- No
- Yes __________________________________________________
- Maybe __________________________________________________

Is there anything else you would like to tell us about the activity or what it has shown you about ChatGPT?

________________________________________________________________

________________________________________________________________

________________________________________________________________

________________________________________________________________

________________________________________________________________

### SMART Goal Grading Rubric

**SMART Goals include all 5 components:**

Specific

Measurable

Attainable

Realistic

Time-bound

**5 pts**

Fully Met

ALL components of a "SMART" goal

**4 pts**

Adequately Met

Four components of a "SMART" goal

**3 pts**

Somewhat Met

Three components of a "SMART" goal

**2 pts**

Somewhat Met

Two components of a "SMART" goal

**1 pt**

Somewhat Met

One component of a "SMART" goal

**0 pts**

Not Met

Zero components of a "SMART" goal
