## Appendix 2 for "Interactive Learning with ChatGPT: Hands-On Practice and Real-Time Feedback in Health Sciences Education for SMART Goal Writing"

### SMART GOALS

WEEK 6.1 LECTURE

| SMART Goals Guide | |
|---|---|
| **Specific** | • What exactly needs to be accomplished?<br>• Who else will be involved?<br>• Where will this take place?<br>• Why do I want to accomplish the goal? |
| **Measurable** | • How will I know I've succeeded?<br>• How much change needs to occur?<br>• How many accomplishments or actions will it take? |
| **Attainable** | • Do I have, or can I get, the resources needed to achieve the goal?<br>• Is the goal a reasonable stretch for me? (neither out of reach nor too easy)<br>• Are the actions I plan to take likely to bring success? |
| **Relevant** | • Is this a worthwhile goal for me right now?<br>• Is it meaningful to me – or just something others think I should do?<br>• Would it delay or prevent me from achieving a more important goal?<br>• Am I willing to commit to achieving this goal? |
| **Time-bound** | • What is the deadline for reaching the goal?<br>• When do I need to take action?<br>• What can I do today? |

Table adapted with permission from ATSU Physical Therapy Goal Writing document.

### GOALS ARE *OUTCOMES*

- Needs to define an **end** state

- Examples:
  - Pt will be taught proper precautions following hip replacement surgery.
  - Pt will be able to demonstrate proper hip replacement precautions during bed mobility, sitting and transfer tasks.

### GOAL QUALITIES

**Concrete** **Measurable** **Testable**

**Predictive**

Examples:

- Pt will demonstrate increased control during reaching movements
- Pt will have good sitting balance.
- Pt will be able to sit unsupported on the edge of the mat table for up to 1 minute without assistance for upper body dressing.

### GOAL FORMAT

- Sentence form
- Table

### CHATGPT EXAMPLE

- Initial Question:

“Create a new patient for physical therapy with subjective findings and specific objective findings of ROM, strength and special tests, palpation; no diagnosis or summary but include normal values.  Can you include specific measurements of ROM and pain in 100 words or less?”

### CHATGPT EXAMPLE

- Prompt:

  “Write a SMART Goal for that patient”

### CHATGPT EXAMPLE

- SMART Goal:

Patient will achieve 90% of normal right shoulder abduction and external rotation ROM (180 degrees abduction, 40 degrees external rotation) within 6 weeks of physical therapy intervention. This will be assessed through goniometric measurements. Additionally, the patient will experience a reduction in pain from 4/10 to 1/10 on the VAS during overhead activities and heavy lifting. Progress will be monitored weekly through objective assessments and pain ratings to ensure steady improvement in shoulder function and pain management.

Response from ChatGPT
based on prompt in prior slide

### HOW CAN WE IMPROVE THIS?

1. Pt will achieve 90% of normal R shoulder ABD & ER AROM in 6 weeks to achieve overhead activities.

2. In 6 weeks, pt will demonstrate reduction in R shoulder pain from 4/10 to 1/10 on VAS during overhead activities and heavy lifting.
