## Appendix 3 for "Interactive Learning with ChatGPT: Hands-On Practice and Real-Time Feedback in Health Sciences Education for SMART Goal Writing"

**Table 1.** Responses to the question, "How do you think ChatGPT can be useful for PTs or PT students?" shown with the number and percentage of student responses per thematic category and representative quotes.

| **Category** | **# (%)** | **Example Response** |
|---|---|---|
| For PT Professional Task (clinical contexts) | 29 (38.16) | "I think it could be helpful with determining how to explain certain activities without non-medical terminology" |
| Knowledge Expansion | 12 (15.79) | "I think that ChatGPT is a good quick resource for students because it generates thoughtful responses and can answer follow up questions as well." |
| Writing Assistance (grammar, organizing thoughts, feedback) | 10 (12.16) | "I believe it would help with proper grammar and punctuation whilst writing a goal as well as organizing a thought." |
| Brainstorming or Considering Alternative Ideas | 9 (11.84) | "It can offer different views to an issue that a person might be having." |
| Unsure or Not Beneficial | 8 (10.53) | "I am not sure since I have never used it" |
| Speed and Efficiency | 6 (7.89) | "Answer questions fast" |
| Creating Study Materials or Practice | 2 (2.63) | "It can help create study tools, and answer questions"<br><br>"It could be interesting to see if they would be a good simulation tool to use for things such as motivational interviewing practice." |

**Table 2.** Responses to the question, "What did you find most helpful about this activity?" shown with the number and percentage of student responses per thematic category and representative quotes.

| Category | # (%) | Example Response |
|---|---|---|
| Virtual Tutor/Feedback | 30 (48.39) | "I found this activity most helpful in getting feedback right then and there after writing the goal instead of waiting around for a professor to grade my work. By the time I would get a grade back from a professor, I am passed caring about what I actually missed on that assignment."<br><br>"ChatGPT not only will give you the answer but it also explains how and why that is the answer." |
| Learning How to Use ChatGPT | 17 (27.42) | "The step by step tutorial regarding using ChatGPT. How to get the correct answer and modifying it in your own words."<br><br>"I got to understand how to use ChatGPT and how it can facilitate learning." |
| Engaging, Hands-On Activity | 10 (16.13) | "making up by own case and scenario and having GPT give me feedback"<br><br>"I think being able to ask the AI to leave out some components of a SMART goal and build your own to then get instant feedback on the edited SMART goal was super useful and I can see myself putting that into use for active recall for studying in different classes."<br><br>"I was able to get a hands on understanding of ChatGPT and SMART goals." |
| Efficiency in Learning | 4 (6.45) | "ChatGPT answers a prompt very fast." |
| Nothing or Not Sure | 1 (1.61) | "NA" |

**Table 3.** Responses to the question, "What did you find least helpful about this activity?" shown with the number and percentage of student responses per thematic category and representative quotes.

| Category | # (%) | Example Response |
|---|---|---|
| ChatGPT Related Dislikes (concerns about using ChatGPT in healthcare or education, inaccurate answers or incomplete answers from ChatGPT) | 16 (28.57) | "That Chat GPT gave too much information and required me to figure out what information was needed and remove the fluff. I also didn't like how the answer wouldn't come in the format we needed for the goal"<br><br>"I think sometimes chat GPT has wrong responses and it makes me weary. I had asked it a question about something and it gave the wrong answer, then when I asked again, it said "sorry for the confusion." |
| Misalignment With Learning Preferences | 12 (21.43) | "I also believe that being taught by Chat GPT is kind of a waste of time because it is something that can be done asynchronously. I chose this PT school because of the in-person learning opportunities, so being taught by AI seems like a waste of money and something I can do on my own time."<br><br>"I felt like there were a lot of reading on the chat portion, which I guess takes place of a lecture, but I don't always learn the best that way." |
| Nothing or Unsure | 10 (17.86) | "I don't think there was anything that was not helpful." |
| Logistical Factors | 9 (16.07) | "The process was long to complete with the technical issues"<br><br>"It is difficult to read prompts GPT writes while many other sounds and voices are happening in the background." |
| Task Wasn't Challenging Enough (too closed nature, wanted more option to explore, be creative, more flexibility) | 9 (16.07) | "I felt like it didn't require a lot of thinking if I didn't want it to. I could've just breezed by." |

**Table 4**. Responses to the question, “Do you have any suggestions about how to improve this activity for future students?” shown with the number and percentage of student responses per thematic category and representative quotes.

| Category | # (%) | Example Response |
|---|---|---|
| Logistics | 19 (39.58) | “The link. Otherwise perfect.”<br><br>“The obvious answer is the WiFi issue. I do think this is helpful and I plan on using it in the future. For reference, after I made an account using my iPhone wifi, I went back onto the school wifi to finish the rest of the actives and it worked fine.” |
| Nothing or Unsure | 17 (35.42) | “no i think it was a good activity and helpful.” |
| Format/Planning (working in groups, doing it outside of class) | 5 (10.42) | “Making it into more of a group activity would be more engaging and fun for the students instead of everyone doing their own. That way people could elaborate more on answers and share ideas.”<br><br>“I think if this were to be implemented in a school setting it could be done asynchronously” |
| More free form exploration of ChatGPT or freely moving through the activity | 4 (8.33) | “Ensure fluidity in moving through the activities.” |
| More examples (case studies, comparing and contrasting how AI writes goals v. professionals) | 2 (4.17) | “Provide actual SMART goals to compare to how the AI writes them to see differences.”<br><br>“more case studies to practice with.” |
| Keep AI Out of Healthcare | 1 (2.08) | “I would say keep AI out of healthcare learning.” |
